# Anxiety levels among Iranian health care workers during the COVID-19 surge: A cross-sectional study

**DOI:** 10.1101/2020.05.02.20089045

**Authors:** Mahbod Kaveh, Fateme Davari-tanha, Shokoh Varaei, Elham Shirali, Nasim Shokouhi, Pershang Nazemi, Mahsa Ghajarzadeh, Elham Feizabad, Mohammad Ali Ashraf

**Affiliations:** Bahrami hospital, Tehran University of Medical Sciences, Tehran, Iran; Yas complex hospital, Tehran University of Medical Sciences, Tehran, Iran; Department of Medical Surgical, School of Nursing and Midwifery, Tehran University of Medical Sciences, Tehran, Iran; Universal Council of Epidemiology (UCE), Universal Scientific Education and Research Network (USERN), Tehran University of Medical Sciences, Tehran, Iran; Department of Obstetrics and Gynecology, Yas complex hospital, Tehran University of Medical Sciences, Tehran, Iran; Student Research Committee, Shiraz University of Medical Sciences, Shiraz, Iran

**Keywords:** COVID-19, Health Care Worker, Anxiety level, Beck Anxiety inventory (BAI), Iran

## Abstract

The recent surge in COVID-19 cases has exposed health care workers (HCWs) to a wide range of psychological stressors and predisposed them to anxiety-related disorders. Here, we investigated the anxiety level in this population. This multi-center, cross-sectional study was performed on 1038 HCWs in 14 hospitals during the COVID-19 pandemic. Beck anxiety inventory (BAI) was used to measure the level of anxiety in this population. In all, 1038 hospital staffs with a mean age of 36.30±8.23 years old participated in this study. Most participants were 31 to 40 years old (43.2), female (87.6%), and nurses (49.5%). The BAI scores for the participants were in a positive skew distribution, with a score range of 0-63, a median of 12 and a mean value of 15.30±11.43. Of the 1038 hospital staff, 411 (39.6%) had moderate to severe anxiety. The anxiety level was significantly higher in health care workers ≤40 years old, women, and nurses. Gender, age, and working positions had the most relation with anxiety, respectively. It seems that HCWs experienced a high level of anxiety in the COVID-19 outbreak. One of the important measures in each epidemic is doing supportive care to maintain the mental well-being of HCWs, especially in higher risk groups, including younger HCWs, women, and nurses.

## 1. Introduction

Since the onset of the coronavirus disease of 2019 (COVID-19) in Wuhan, it has spread to about 212 countries/territories and caused more than 230000 deaths worldwide. The emergence of the disease caused a large burden on hospitals and health care workers (Remuzzi and Remuzzi, 2020). This increased the need for health care workers and health care resources. However, there an increasing need for psychological support as well (Chen et al., 2020).

Health care workers (HCWs) are one of the most stressed occupations in this outbreak (Sim, 2020). The recent surge in COVID-19 cases has exposed them to a wide range of psychological stressors. Extended working hours and the shortage of personal protection equipment (PPE) has made their mental health more vulnerable than ever (Ayanian, 2020). HCWs’ current situation predisposes them to anxiety and depression-related disorders, which might lead to poor performance and disability in decision making (Kang et al., 2020).

Health care workers are in the frontline in the battle with COVID-19, and their mental and physical health should be a priority (Tsamakis et al., 2020). Therefore, there is an increasing need to recognize the most stressed hospital workers and provide more support for this group.

Beck Anxiety Inventory (BAI) is a valid self-report questionnaire for measuring anxiety levels in the general population (Julian, 2011; Osman et al., 1997). We used the validated Persian version of BAI to measure the rate of anxiety-related symptoms and the level of anxiety among our health care workers (Hossein Kaviani and Mousavi, 2008).

This multi-center study was aimed to evaluate the level of anxiety in health care workers of tertiary care hospitals designated for COVID-19 patients. This data can help us recognize the most vulnerable groups in health care workers of Tehran hospitals during the covid-19 pandemic. To our knowledge, this study is the first report among Iranian HCWs during the COVID-19 pandemic.

## 2. Methods

### 2.1. Participants and study overview

This cross-sectional, multi-center, web-based survey was started on March 25, 2020, and 1038 health care workers from 14 tertiary care hospitals affiliated with Tehran University of Medical Sciences (TUMS) participated in the study. The questionnaire was made online and was sent to all 14 designated hospital staff through social media. The staff could decide whether or not they want to participate in the survey. Respondents were chosen from the doctors, nurses, midwives, and other staff who were working in the designated COVID-19 wards. Other personnel who did not have exposure to the patients and other hospital employees who were working outside patient areas were excluded from the study. All participants consented by filling the questionnaire form. The confidentiality of their name and personal information was assured, and the principle of confidentiality was observed. The study protocol was approved by the Tehran University of Medical Sciences ethics committee (IR.TUMS.VCR.REC.1399.087).

### 2.2. Data collection

Basic demographic data, including age, gender, and their working position was asked through a web-based questionnaire. Their basic information and working positions were then confirmed by originating hospitals. The level of anxiety was measured using the validated Persian version of 21-question Beck Anxiety Inventory (BAI), and each question was graded from 0 to 3 (0=minimal; 1=mild, 2=moderate; 3=severe) (Hossein Kaviani and Mousavi, 2008). In this survey, the subjects were asked whether they had common symptoms of anxiety over the past working week. The sums of all questions score were defined as an indicator of anxiety, which provides a score between 0 to 63 (0 to 7= minimal anxiety, 8 to 15= mild anxiety, 16 to 25= moderate anxiety, and 26 to 63= severe anxiety). Only a single response to the questionnaire was permitted for each person. The survey was continued up to March 31, 2020.

### 2.3. Outcomes

The participants were categorized twice in terms of working positions. Once classified as two groups of first-line workers vs. second-line workers, and next classified as four groups of physicians, nurses, midwives, and other working positions. Other working positions consisted of paramedics, nurse assistants, and administrative officers. First-line workers were defined as the group of personnel who have direct exposure to confirmed/suspected admitted COVID-19 patients and who were involved in aerosol-generating procedures (e.g., tracheal intubation, non-invasive ventilation, tracheotomy, cardiopulmonary resuscitation, manual ventilation before intubation, bronchoscopy) (Organization, 2020). Second-line workers were defined as other personnel who did not have direct exposure with the patients, and those who were not involved in aerosol-generating procedures. Demographic data including age (20-25, 26-30, 31-40, 41-50, or >50 years), gender (male or female), and working positions (physicians, nurses, midwives, other positions / front-line vs. second-line workers) were collected in the study population. The overall score of anxiety was reported as a four-level variable (minimal anxiety, mild anxiety, moderate anxiety, severe anxiety) in our study population. Also, the association of anxiety level and age, gender, or different working positions were investigated.

### 2.4. Statistical analysis

Data analysis was performed at both descriptive and inferential levels. At the descriptive level, mean± standard deviation was used for interval variables, frequency, and percent for categorical variables were used. At the inferential level, the Kolmogorov-Smirnov test was used to evaluate the distribution of the data. Pearson correlation coefficient test was used to investigate the relation between anxiety level and the age of the population. Also, One-way analysis of variance was used to investigate the relation between the level of anxiety and gender, age groups, working positions (with respect to both grouping [physicians, nurses, midwives, others vs. first-line and second-line workers]) of our participants. In addition, Tukey’s post-hoc test was used to examine the difference in anxiety levels between different ages and working positions groups. P-value<0.05 was considered significant, and 95 % confidence interval was considered acceptable. All data were analyzed using SPSS software, version 24.0 (IBM).

## 3. Results

In total, 1038 health care workers from 14 designated hospitals participated in this study. The mean age of the participants was 36.30±8.23 years old (range, 20 to 66). Most participants were women (909 [87.6%]) and aged between 31 to 40 years (449 [43.2%]). In this study, participants were categorized twice in terms of working positions. Once classified as two groups of front-line workers (933[89.9%]) vs. second-line workers (105[10.1%]), and next classified as four groups of nurses (514[49.5%]), physicians (214[20.6%]), midwives (143[13.8%]), and others (167[16.1%]). Other working positions consisted of paramedics, nurse assistants, and administrative officers. (as shown in table 1)

**Table 1.** Basic Demographic data of participants.

| Demographic data | N(%) |
| --- | --- |
| **Age** |  |
| 20-25 years | 89 (8.6) |
| 26-30 years | 193 (18.6) |
| 31-40 years | 449 (43.2) |
| 41-50 years | 217 (20.9) |
| ≥51 years | 56 (5.4) |
| Missing | 34 (3.3) |
| **Gender** |  |
| Female | 909 (87.6) |
| male | 129 (12.4) |
| **Job type** |  |
| **First line** | 933 (89.9) |
| **Second line** | 105 (10.1) |
| **Occupation** |  |
| physician | 214 (20.6) |
| Nurse | 514 (49.5) |
| Midwife | 143 (13.8) |
| Others<sup>a</sup> | 167 (16.1) |

<sup>a</sup> Including paramedics, nurse assistants and administrative officers.

The scores of BAI for the participants had positive skew distribution, which means the majority of health care workers had minimal to mild anxiety (N = 627, 60.4%). The other 411 workers had moderate to severe anxiety (39.6%). Our population’s overall BAI score ranged from 0 to 63 and had a mean value of 15.30±11.43 with a median of 12.

The score of anxiety was higher among employees less than 40 years old. The age groups under 40 years had a similar mean anxiety score (age 20-25: mean anxiety score of 16.46; age 26-30: mean anxiety score of 16.33; age 31-40; mean anxiety score of 16.16). However, anxiety score was lower in ≥41 years old workers, and the mean score was similar among age groups above 40 years (age 41-50: mean anxiety score of 12.73; age ≥51: mean anxiety score of 12.82). Male workers had lower anxiety than females (mean anxiety score: 10.26 vs. 16.03). Male employees between 31-40 years old had the lowest anxiety (mean score of 9.05). However, the highest anxiety was seen in female employees between 31-40 years of age (mean score of 17.19). Analysis of different working positions shows the lowest anxiety score in physicians (mean score of 13.03) followed by midwives, nurses, and others (mean score of 15.80 vs. 15.88 vs. 16.01, respectively). The highest anxiety was seen in female workers of other working positions group (mean score of 16.82). Table 2 presents the BAI score (mean ± standard deviation) in male and female workers according to their age and working positions.

**Table 2.** Mean and standard deviation of BAI scores in male and female workers according to their age and working positions.

| Age groups |  | Mean (S.D) |  |  |  |  |
| --- | --- | --- | --- | --- | --- | --- |
|  |  | 20-25 | 26-30 | 31-40 | 41-50 | > 51 |
| Sex |  |  |  |  |  |  |
| Female |  | 16.56 (12.25) | 16.63 (11.63) | 17.19 (11.92) | 13.29 (10.10) | 14.17 (10.30) |
| Male |  | 14.25 (9.36) | 14.42 (12.19) | 9.05 (8.01) | 9.14 (9.08) | 6.60 (3.63) |
| Total |  | 16.46 (12.10) | 16.33 (11.70) | 16.16 (11.81) | 12.73 (10.05) | 12.82 (9.87) |
| Work positions |  | Mean (S.D) |  |  |  |  |
|  |  | Physician | Nurse | Midwife | Others | Total |
| Sex |  |  |  |  |  |  |
| Female |  | 14.51 (11.35) | 16.37 (11.79) | 15.89 (10.00) | 16.82 (12.16) | 16.03 (11.51) |
| Male |  | 7.92 (5.48) | 10.40 (9.70) | 3.00 (-) | 13.26 (12.12) | 10.26 (9.42) |
| Total |  | 13.03 (10.68) | 15.88 (11.74) | 15.80 (10.02) | 16.01 (12.21) | 15.30 (11.43) |

As shown in table 3, One-way analysis of variance was used to examine the association between age, sex, and working positions variables and the anxiety score. The level of anxiety was significantly different in male and female workers (F (1, 1035) = 29.45, P= 0.0001), and according to table 2, the level of anxiety was higher in female workers. There was a statistically significant difference in the level of anxiety in different age groups (F (4, 999) = 4.71, P= 0.001). Pearson correlation coefficient test showed a negative relation between the age and the level of anxiety (R = 0.13, P = 0.0001). Higher ages were associated with lower anxiety in our population. There was not a relation between first-line/second-line workers and anxiety (F (1, 1035) = 0.05, P=0.82). However, different working positions (physicians, nurses, midwives, others) had a different level of anxiety, which was statistically significant (F (3, 1033) = 3.59, P = 0.01). According to partial eta squared, it can be concluded that sex, age, and four groups of working positions had the most relation with anxiety, respectively.

**Table 3.**
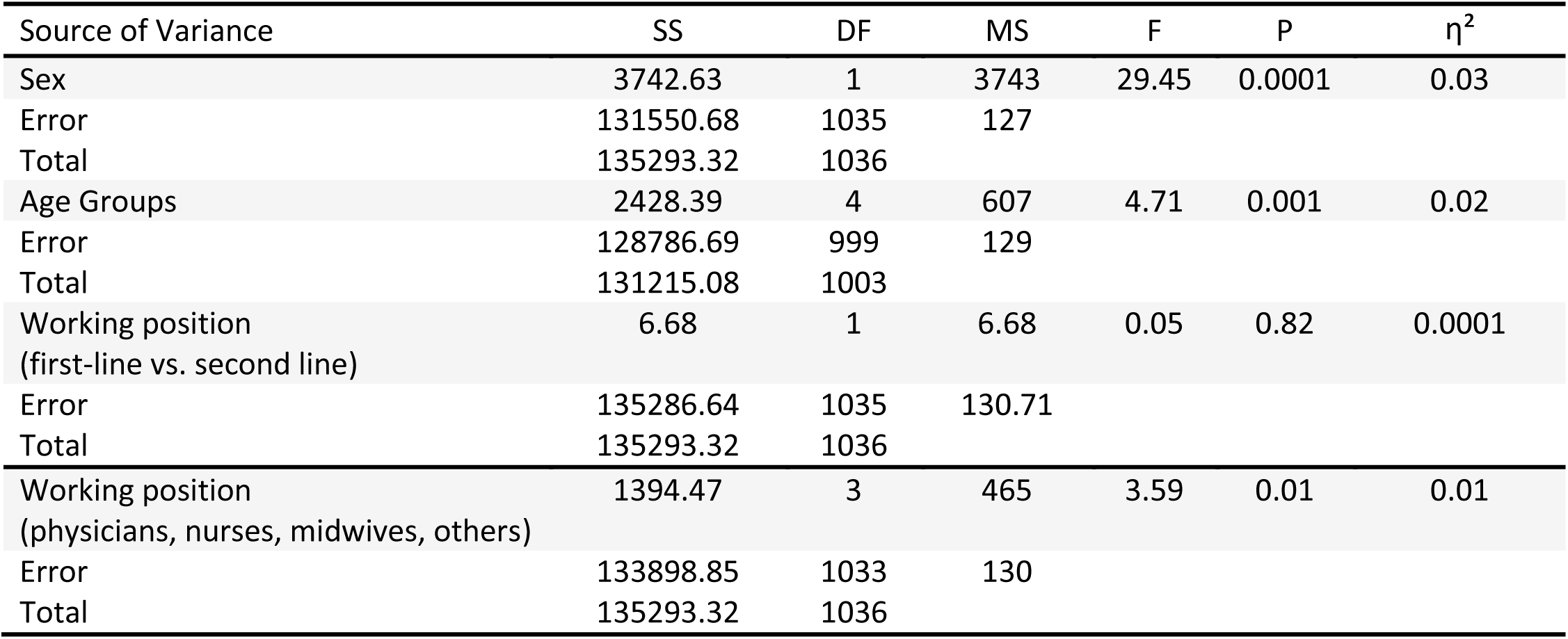
The results of One way analysis of variance to examine the association of demographic variables and anxiety scores

| Source of Variance | SS | DF | MS | F | P | $\eta^2$ |
| --- | --- | --- | --- | --- | --- | --- |
| Sex | 3742.63 | 1 | 3743 | 29.45 | 0.0001 | 0.03 |
| Error | 131550.68 | 1035 | 127 |  |  |  |
| Total | 135293.32 | 1036 |  |  |  |  |
| Age Groups | 2428.39 | 4 | 607 | 4.71 | 0.001 | 0.02 |
| Error | 128786.69 | 999 | 129 |  |  |  |
| Total | 131215.08 | 1003 |  |  |  |  |
| Working position<br>(first-line vs. second line) | 6.68 | 1 | 6.68 | 0.05 | 0.82 | 0.0001 |
| Error | 135286.64 | 1035 | 130.71 |  |  |  |
| Total | 135293.32 | 1036 |  |  |  |  |
| Working position<br>(physicians, nurses, midwives, others) | 1394.47 | 3 | 465 | 3.59 | 0.01 | 0.01 |
| Error | 133898.85 | 1033 | 130 |  |  |  |
| Total | 135293.32 | 1036 |  |  |  |  |

The results of the analysis of variance were followed by Tukey’s post-hoc test to examine the difference between different ages and working positions. As shown in table 4, the age group of 41-50 years had lower anxiety than 26-30 and 31-40 year groups. Other age groups were similar in terms of the level of anxiety. Furthermore, the analysis showed that nurses had higher anxiety than physicians. However, there was not a statistically significant difference between other working positions groups (according to table 4).

**Table 4.**
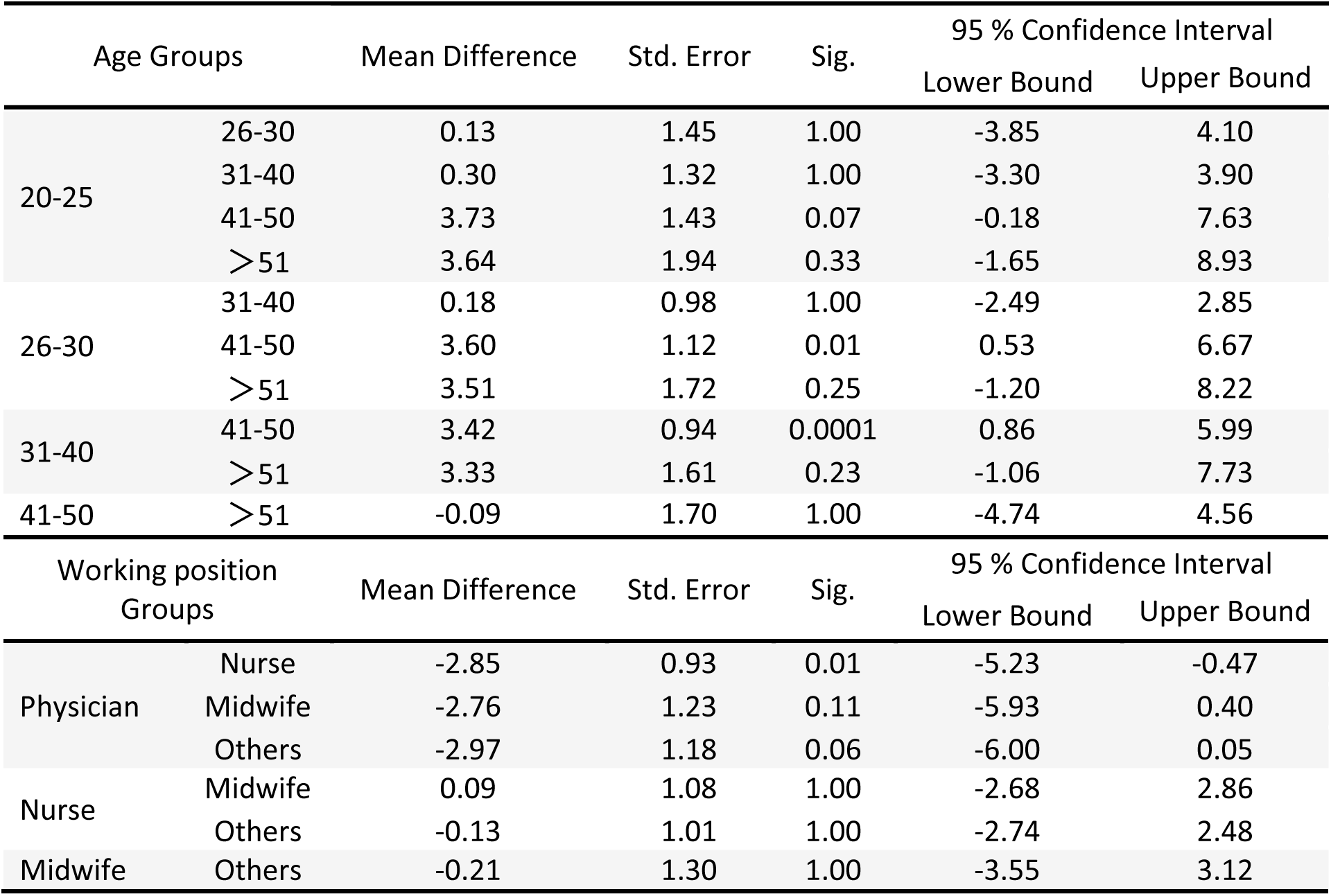
Tukey’s post-hoc test for comparison of anxiety scores between age and four working positions groups.

BAI is a 21-item questionnaire, and each item scores between 0 to 3 based on the severity. Assessing each item gives us valuable information. As shown in figure 1, the following items had a mean score above 1: 1. fear of worst happening, 2. unable to relax, 3. unsteady, 4. nervous, 5. Scared. On the other hand, five of the other items had a mean score below 0.4 as follows: 1. Faint/lightheaded, 2. Face flushed, 3. Shaky/unsteady, 4. Hands trembling, 5. Wobbliness in legs. The items had a lower mean score in the physicians’ group, and a higher mean score in nurses and midwives (as shown in figure 1).

**Figure 1.**
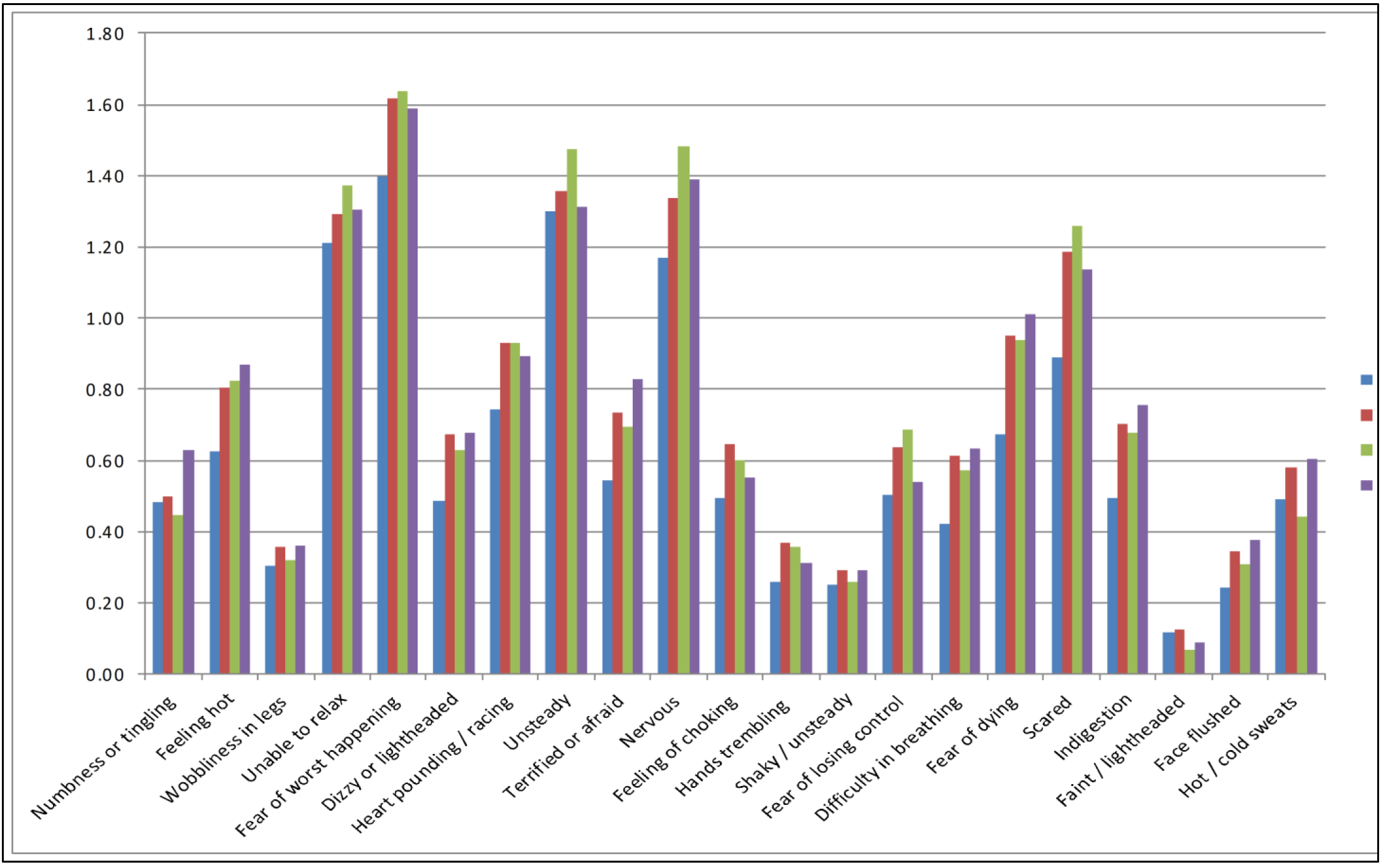
Mean score of each Beck Anxiety Inventory items according to participants working positions.

## 4. Discussion

This multi-center study was aimed to evaluate the level of anxiety among health care workers at Tehran tertiary hospitals designated to care for COVID-19 patients. The results can be an indicator of anxiety among Iranian HCWs. We identified the most at-risk group in terms of psychological stress and anxiety among hospital personnel amid the COVID-19 pandemic.

Higher anxiety levels were found in female health care workers who were less than 40 years of age. Lowest anxiety was seen in physicians, followed by midwives, nurses, and other working positions, respectively. These findings were consistent with previous studies, in which nurses and women were found to have more severe anxiety symptoms (Huang et al., 2020; Lai et al., 2020).

In our study, lower ages were associated with higher levels of anxiety. This might be associated with higher educational levels and higher work experience of older staff in high-stress hospital environments, which can lead to higher understanding of Covid-19 and knowing how to effectively use personal protection equipment (PPE). This finding is consistent with Øyane et al. study, who found a negative association between years of work experience and anxiety in Norwegian nurses (Øyane et al., 2013).

Beck anxiety inventory was used to evaluate the level of anxiety among our population in Tehran hospitals. Kaviani et al. showed that the Persian version of BAI has good psychometric properties in evaluating anxiety levels among the Iranian population (Hossein Kaviani and Mousavi, 2008).

In total, 18.1% of participants experienced severe anxiety, 21.5% moderate anxiety, and 31.8% mild anxiety. Overall, about 40% of all participants experienced moderate to severe anxiety level, which was in accordance with Lai et al. study, who reported anxiety in 44.6 % of HCWs of 34 hospitals in China during the COVID-19 epidemic (Lai et al., 2020). Also, in SARS and H1N1 influenza epidemic, a significant proportion of HCWs displayed high anxiety levels (Chua et al., 2004; Goulia et al., 2010; Tam et al., 2004). In total, 71.4% of HCWs had some degree of anxiety, which is similar to previous studies about HCWs during the SARS and MERS epidemics. Alsahafi et al. reported 61.2% of HCWs had anxiety about contracting MERS-CoV (Alsahafi et al., 2016). Koh et al. reported that 56% of hospital staff had increased work stress during the SARS epidemic (Koh et al., 2005).

Although the psychological response of HCWs to an outbreak is complicated, the reason why young people and people with lower intermediate technical titles had higher anxiety can be due to their lack of control and authority in their work and fear of changes in their work. Other sources of distress may include fear of contagion, feelings of vulnerability, concerns about the health of themselves and their family, perceived stigma, and being isolated (Maunder et al., 2006). Therefore, these factors might be present in our population, as well, and should be addressed.

Anxiety levels were higher in nurses in comparison to physicians. This can be due to physicians’ higher educational levels and information on the disease. Another explanation might be due to greater patient contact by nurses, greater risk perception, longer working hours, and lack of adequate PPE (Brooks et al., 2018; Lai et al., 2020).

COVID-19 surge caused a new situation, which demands an excessive load of work by health care personnel. It is challenging for HCWs to confront the new situation and predispose them to anxiety-related disorders. By recognizing the most vulnerable HCWs, it is essential to provide more support for this group to cope with the emerging situation. Some factors, including social appreciation, greater family/friends support, hospital psychological support, providing adequate protective equipment, and personnel training, can be helpful for coping with this situation (Brooks et al., 2018; Su et al., 2007).

This study had some limitations. First, we only evaluated the anxiety level of HCWs and not other psychological disorders. Second, the study was done in a short period (6 days), and about one month after the onset of the COVID-19 pandemic in Iran, while the anxiety level of HCWs may vary at different times during the epidemic. Currently, little research has been done on this topic, and we had not been able to compare the results of our study extensively with previous studies.

## 5. Conclusion

It seems that Iranian HCWs experienced a high level of anxiety in the COVID-19 outbreak. One of the important measures in each epidemic is doing supportive care to maintain the mental well-being of HCWs, especially in higher-risk groups, including younger HCWs, women, and nurses.

## Data Availability

the data regarding the manuscript will be available as reasonable request.

## Acknowledgements

The authors would like to acknowledge the hospital stuff who participated in this survey.

